# MORPHOMETRIC IMAGE ANALYSIS COMPARISON OF BASAL CELL CARCINOMAS & MELANOMAS OF THE HEAD & NECK VERSUS OTHER SITES

**DOI:** 10.1101/2024.01.05.24300900

**Authors:** Jonas Hue, Jinendra Ekanayake, Jamshid Dehmeshki, Jagtar Dhanda

## Abstract

**Background:** Management of skin cancers are heavily dependent on clinical diagnosis via dermoscopy. Dermoscopic morphology of basal cell carcinomas (BCC) and melanomas influence diagnosis and may be influenced by anatomic location.

**Objective:** In this study, we aimed to investigate the morphologic differences between anatomic sites of the head and neck (H&N) versus other body sites in basal cell carcinomas and melanomas.

**Patients and methods:** The publicly available HAM10000 dataset was used in this study. Morphometric image analysis of the BCCs (n=422) and melanomas (n=868) in this dataset was performed using an open-source image analysis software. Univariate and multivariate statistical analysis was done to identify differences between H&N and other anatomic sites. The multifactorial data was further interrogated with dimensionality reduction techniques, linear discriminant analysis, principal component analysis and t-distributed stochastic neighbour embedding.

**Results:** Univariate and multivariate statistical analyses revealed significant differences between H&N and other sites for both BCCs and melanomas (*P*<0.05). Fifty-three univariate and 11 multivariate features were found to be statistically significant in the BCC group. Thirteen univariate and 8 multivariate features were statistically significant in the melanoma group. Dimensionality reduction via linear discriminant analysis of the BCC groups revealed modest separation of the data by anatomical site. However, melanomas appeared to be more homogenous across H&N and other body sites.

**Conclusion:** BCCs of the H&N may be morphologically different to BCCs of other body sites. This may influence the accuracy of computer-assisted diagnostic tools and specialist clinicians working predominantly in the H&N region should exercise caution when employing these tools in clinical practice.

## Introduction

Basal cell carcinoma (BCC) is the most common skin cancer^1^, creating a significant burden on healthcare services. Fortunately, metastasis is rare and these lesions can often be treated with local surgical and non-surgical therapies^2^. Treatment protocols and surgical margins are influenced by clinical diagnosis and risk assessment (high versus low) as certain BCC subtypes and small lesions may be definitively treated without a prior biopsy^3^.

Cutaneous melanoma is the third most common skin cancer but responsible for the most number of skin cancer deaths due to metastatic disease^4^. Early diagnosis is particularly crucial to ensure better patient outcomes due to the great propensity for spread in this type of cancer^5^.

Clinicians employ various tools to achieve accurate diagnoses, such as dermoscopy—a non-invasive epiluminescence microscopy technique—and, more recently, artificial intelligence (AI). The development of AI models for assisting in the clinical diagnosis of skin lesions from dermoscopic images has garnered significant interest, with many models demonstrating promising outcomes. These image-based AI models often utilise morphometric features, among other characteristics, of skin lesions to reach a diagnosis. However, a notable challenge is that many models operate as black boxes, lacking internal source code, which complicates the understanding of the decision-making algorithms and the features involved^6^. The absence of transparency raises difficulties in comprehending the factors influencing the model’s decisions. Theoretically, variations in morphology could impact classifier accuracy based on the model inputs. To our knowledge, there has been a lack of comprehensive analyses comparing the morphologic heterogeneity of malignant skin lesions across different anatomical sites.

Anatomic location of a skin lesion influences how it is treated. Skin malignancies of the head and neck (H&N) in particular may present to and be treated by various clinicians such as general practitioners, dermatologists, plastic surgeons and oral and maxillofacial surgeons^2^. These clinical specialities receive different training ^7–9^ and a dermoscopic AI model may help in diagnostic equity and level the playing field. However, such a model must be validated on external datasets and perform well on skin lesions from different sites.

Anatomical site of the H&N is also a point of consideration when planning treatment due to the underlying anatomy, difficulty of surgery and cosmetic considerations. Furthermore, these underlying anatomic variations may contribute to a difference in the behaviour and growth of skin cancers^10^, resulting in quantifiable changes in their surface morphology. The head and neck region is also a high risk site due to sun exposure, an aetiological factor for cutaneous malignancies. Sun exposure has been shown to be associated with certain subtypes of BCCs and may have an influence on their histopathology^11^, which will in turn, affect their dermoscopic features^12^.

The aim of this study is to investigate and analyse the morphometric differences in skin lesions, specifically focusing on Basal Cell Carcinomas (BCCs) and melanomas, with a particular emphasis on those located in the H&N region compared to lesions in other anatomical sites.

We hypothesize that there are distinct and quantifiable morphometric variations between BCCs and melanomas situated in the H&N region, in contrast to lesions found in other anatomical sites associated with each respective type of cancer.

## Methods

In this study, we used the publicly available HAM10000 dataset^13^. Lesions that were not imaged in their entirety (touching the borders of the image) were excluded. BCCs (n=422) and melanomas (n=868) were identified and split into two groups (Head & Neck versus Others) for each cancer type. This resulted in a total of 112 BCCs and 113 melanomas from the H&N. On the other hand, there were 310 BCCs and 755 melanomas from other sites (Table 1).

**Table 1.** Clinical features of the patient cohort.

|  |  | **BCC**<br>**(n=422)** | **Melanoma**<br>**(n=868)** |
| --- | --- | --- | --- |
| **Age(mean/range)** |  | 66.2 (20-85) | 59.6 (20-85) |
| Sex | **Male, n (%)** | 259 (61.4) | 540 (62.2) |
|  | **Female, n (%)** | 163 (38.6) | 328 (37.8) |
| Site | **H&N, n (%)** | 112 (26.5) | 113 (13.0) |
|  | **Other Site , n (%)** | 310 (73.5) | 755 (87.0) |

Morphometric image analysis was performed on the segmented binary masks^14^ using the open-source software, CellProfiler^15^ (version 4.2.1). Each dermoscopic skin lesion was identified as an individual object. A total of 98 morphometric features were quantified with the MeasureAreaShape module, including their eccentricity, solidity, form factor and image moments. Elaboration and explanation of each individual feature can be found in the CellProfiler version 4.2.1 Manual. The two available clinicopathological features of age and sex were also tested for significance.

Statistical analysis was performed in R using the “stats” package. Univariate analysis was performed using the Wilcoxon’s rank-sum test due to its robustness in handling non-parametric data. Logistic regression was used for multivariate analysis. For sex-based analysis, Pearson’s Chi-Squared test was used for univariate assessment. Statistical significance accepted at a threshold of *P*< 0.05. These methods were selected based on their appropriateness for the data type and research objectives.

Dimensionality reduction and data visualisation techniques were employed to further interrogate the data. Linear discriminant analysis, principal component analysis (PCA) and t-distributed Stochastic Neighbour Embedding (t-SNE) were performed in R with the “MASS” and “Rtsne” packages.

## Results

### BCCs of the head & neck exhibit moderate morphological differences to BCCs of other body sites

Uni- and multivariate statistical analysis of the morphometric features quantified of BCCs, revealed several statistically significant differences between H&N BCCs and other sites. In total, 53 variables were statistically significant on univariate analysis (Table 2). Eleven variables were found to be statistically significant with multivariable logistic regression (Table 3). The McFadden’ R2 for the logistic regression model of 0.313 showed an excellent fit of the data (a range of 0.2-0.4 indicates an excellent fit). In general, H&N BCCs had higher solidity (*P*=0.028) and form factor (*P*=0.001), and lower compactness (*P*=0.001) (Figure 1), indicating a more rounded shape with less irregular borders.

**Figure 1.**
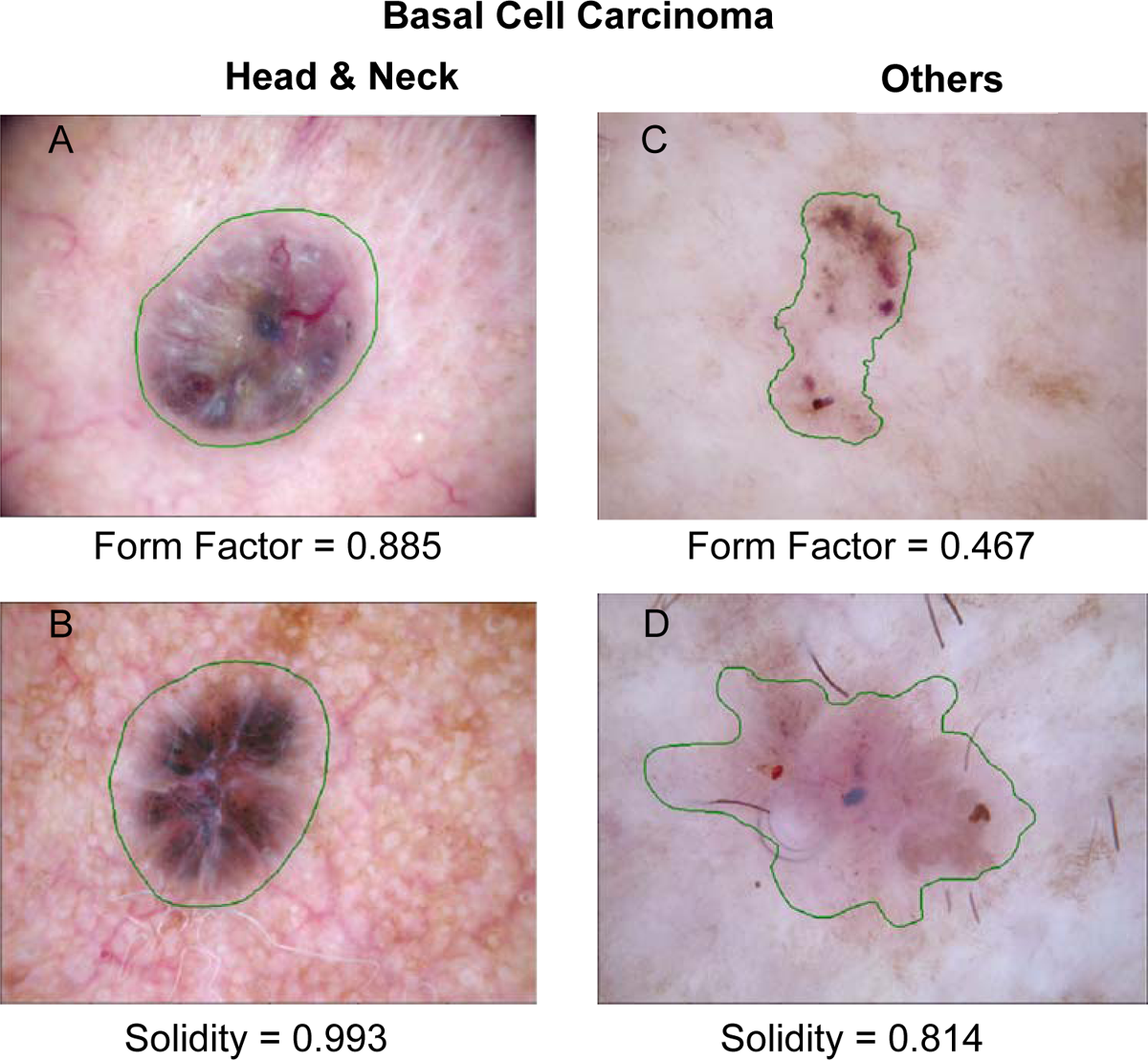
Example BCCs of the H&N exhibiting quantifiable morphological differences to BCCs of other body sites. (A) and (B) showing BCCs of the H&N with higher form factor and solidity compared to (C) and (D) showing BCCs of other sites with less rounded borders.

**Table 2.** Statistically significant BCC features on univariate Wilcoxon’s rank sum test. Statistical significance accepted at *P*<0.05.

|  | **H&N**<br>**Median** | **H&N IQR** | **Others**<br>**Median** | **Others**<br>**IQR** | **Univariate**<br>**Wilcoxon's**<br>**Rank Sum**<br>**Test $P$** |
| --- | --- | --- | --- | --- | --- |
| Area | 42474 | 46583 | 57386 | 51866 | 0.001 |
| Bounding Box Area | 58437 | 70875 | 80122 | 80635 | 0.002 |
| Central Moment 0,0 | 42474 | 46583 | 57386 | 51866 | 0.001 |
| Central Moment 0,2 | 1.65E+08 | 4.18E+08 | 3.09E+08 | 6.66E+08 | 0.001 |
| Central Moment 1,1 | -2.35E+06 | 4.56E+07 | 2.16E+05 | 5.93E+07 | 0.008 |
| Central Moment 1,3 | -5.87E+09 | 5.12E+11 | 8.71E+08 | 6.29E+11 | 0.008 |
| Central Moment 2,0 | 1.40E+08 | 3.06E+08 | 2.56E+08 | 4.42E+08 | 0.002 |
| Central Moment 2,2 | 4.46E+11 | 1.63E+12 | 9.83E+11 | 3.34E+12 | 0.001 |
| Compactness | 1.24 | 0.16 | 1.29 | 0.19 | 0.001 |
| Convex Area | 44420 | 48016 | 58953 | 57518 | 0.001 |
| Equivalent Diameter | 232.5 | 127.0 | 270.3 | 124.2 | 0.001 |
| Form Factor | 0.804 | 0.102 | 0.776 | 0.112 | 0.001 |
| Hu Moment 0 | 0.168 | 0.010 | 0.170 | 0.014 | 0.039 |
| Inertia Tensor Eigen<br>values 0 | 4571 | 5472 | 6339 | 6282 | 0.001 |
| Inertia Tensor Eigen<br>values 1 | 2469 | 2823 | 3413 | 3268 | 0.002 |
| Inertia Tensor 0,0 | 3864 | 4831 | 5409 | 5939 | 0.001 |
| InertiaTensor 0,1 | 146.4 | 846.0 | -26.4 | 1140.4 | 0.009 |
| InertiaTensor 1,0 | 146.4 | 846.0 | -26.4 | 1140.4 | 0.009 |
| InertiaTensor 1,1 | 3045 | 3157 | 4079 | 3937 | 0.006 |
| Major Axis Length | 270.4 | 161.9 | 318.5 | 158.5 | 0.001 |
| Max Feret Diameter | 279.9 | 172.5 | 325.4 | 164.5 | 0.001 |
| Maximum Radius | 95.4 | 50.9 | 110.1 | 50.9 | 0.003 |
| Mean Radius | 34.9 | 17.8 | 39.7 | 17.4 | 0.003 |
| Median Radius | 31.2 | 15.7 | 35.8 | 15.9 | 0.003 |
| Min Feret Diameter | 200.1 | 115.5 | 237.0 | 118.4 | 0.001 |
| Minor Axis Length | 198.8 | 110.2 | 233.7 | 113.1 | 0.002 |
| Normalized Moment 0,2 | 0.092 | 0.027 | 0.099 | 0.031 | 0.018 |
| Normalized Moment 1,1 | -5.59E-03 | 2.45E-02 | 9.48E-04 | 2.22E-02 | 0.010 |
| Normalized Moment 1,3 | -8.05E-04 | 4.20E-03 | 2.17E-04 | 4.14E-03 | 0.009 |
| Normalized Moment 2,0 | 0.074 | 0.024 | 0.070 | 0.021 | 0.029 |
| Normalized Moment 3,1 | -8.74E-04 | 3.92E-03 | 1.51E-04 | 3.15E-03 | 0.016 |
| Normalized Moment 3,3 | -1.18E-04 | 5.10E-04 | 2.13E-05 | 5.02E-04 | 0.014 |
| Orientation | -33.1 | 109.4 | 5.3 | 139.7 | 0.013 |
| Perimeter | 820.1 | 495.4 | 974.2 | 525.8 | 0.001 |
| Solidity | 0.970 | 0.036 | 0.964 | 0.036 | 0.028 |
| Spatial Moment 0,0 | 42474 | 46583 | 57386 | 51866 | 0.001 |
| Spatial Moment 0,1 | 5.21E+06 | 9.69E+06 | 8.57E+06 | 1.32E+07 | 0.001 |
| Spatial Moment 0,2 | 7.91E+08 | 2.21E+09 | 1.62E+09 | 3.46E+09 | 0.001 |
| Spatial Moment 0,3 | 1.40E+11 | 5.25E+11 | 3.43E+11 | 9.63E+11 | 0.001 |
| Spatial Moment 1,0 | 4.72E+06 | 8.11E+06 | 7.89E+06 | 1.07E+07 | 0.001 |
| Spatial Moment 1,1 | 6.02E+08 | 1.49E+09 | 1.10E+09 | 2.26E+09 | 0.001 |
| Spatial Moment 1,2 | 9.63E+10 | 3.18E+11 | 2.09E+11 | 5.61E+11 | <0.001 |
| Spatial Moment 1,3 | 1.71E+13 | 8.23E+13 | 4.35E+13 | 1.62E+14 | <0.001 |
| Spatial Moment 2,0 | 6.58E+08 | 1.62E+09 | 1.28E+09 | 2.41E+09 | 0.002 |
| Spatial Moment 2,1 | 8.79E+10 | 2.70E+11 | 1.86E+11 | 4.60E+11 | 0.001 |
| Spatial Moment 2,2 | 1.29E+13 | 5.91E+13 | 3.39E+13 | 1.08E+14 | <0.001 |
| Spatial Moment 2,3 | 2.28E+15 | 1.32E+16 | 6.99E+15 | 2.96E+16 | <0.001 |
| Zernike 0,0 | 0.717 | 0.126 | 0.685 | 0.133 | 0.024 |
| Zernike 2,0 | 0.162 | 0.030 | 0.167 | 0.028 | 0.017 |
| Zernike 4,0 | 0.045 | 0.033 | 0.036 | 0.035 | 0.009 |
| Zernike 8,4 | 6.42E-03 | 5.53E-03 | 7.35E-03 | 5.97E-03 | 0.018 |
| Zernike 9,7 | 3.40E-03 | 2.73E-03 | 4.04E-03 | 3.19E-03 | 0.041 |
| Age | 70.0 | 15.0 | 67.5 | 20.0 | <0.001 |

**Table 3.**
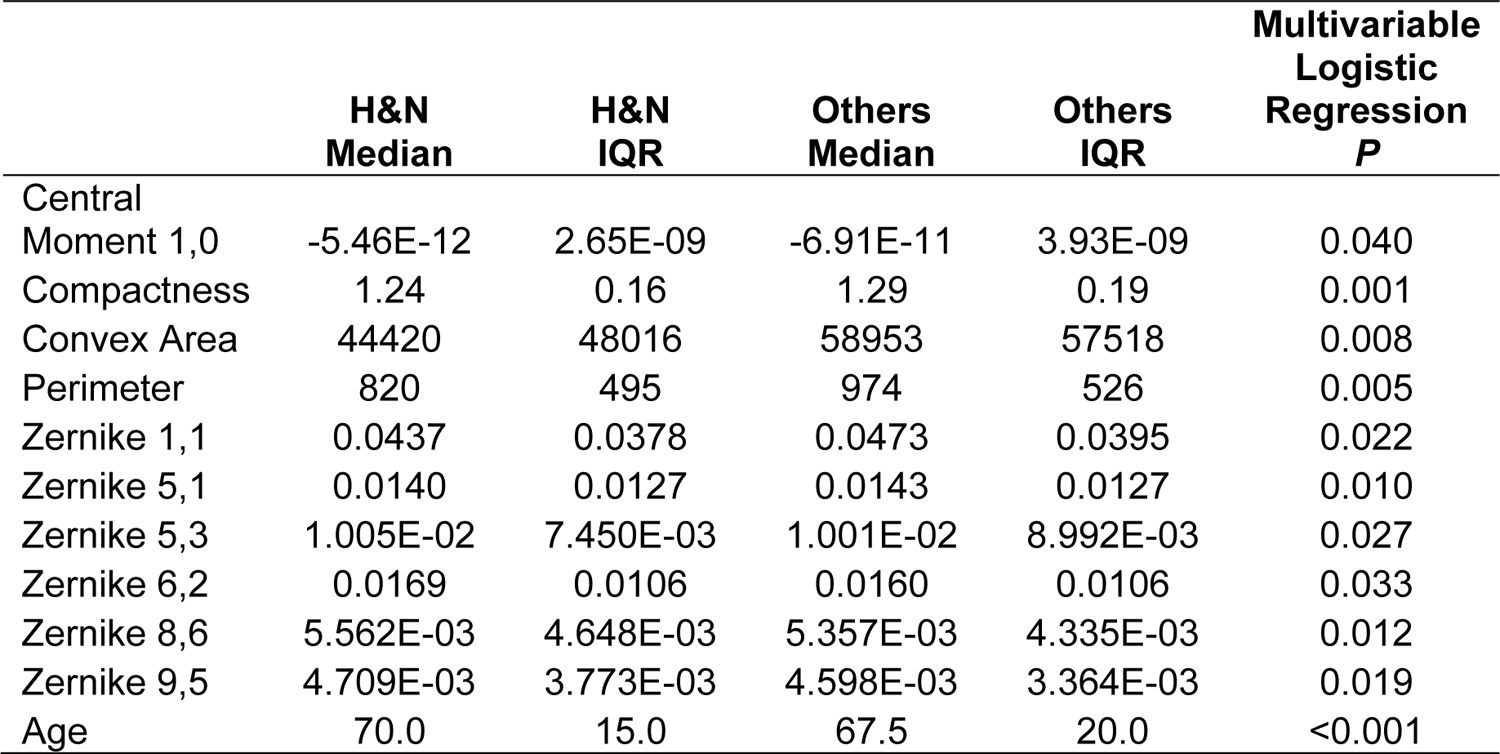
Statistically significant BCC features on multivariable logistic regression. Statistical significance accepted at *P*<0.05.

Unsurprisingly, upon further interrogation of the data with various dimensionality reduction methods, we see significant overlap between H&N BCCs and other sites. Despite their morphological differences, they are still the same type of cancer and we would expect many of them to look similar. PCA and t-SNE plots did not reveal any separation of the data based on their anatomical labels (Figure 2A and 2B). However, the LDA histogram showed moderate separation of the data with the H&N group shifted to the left (Figure 2C).

**Figure 2.**
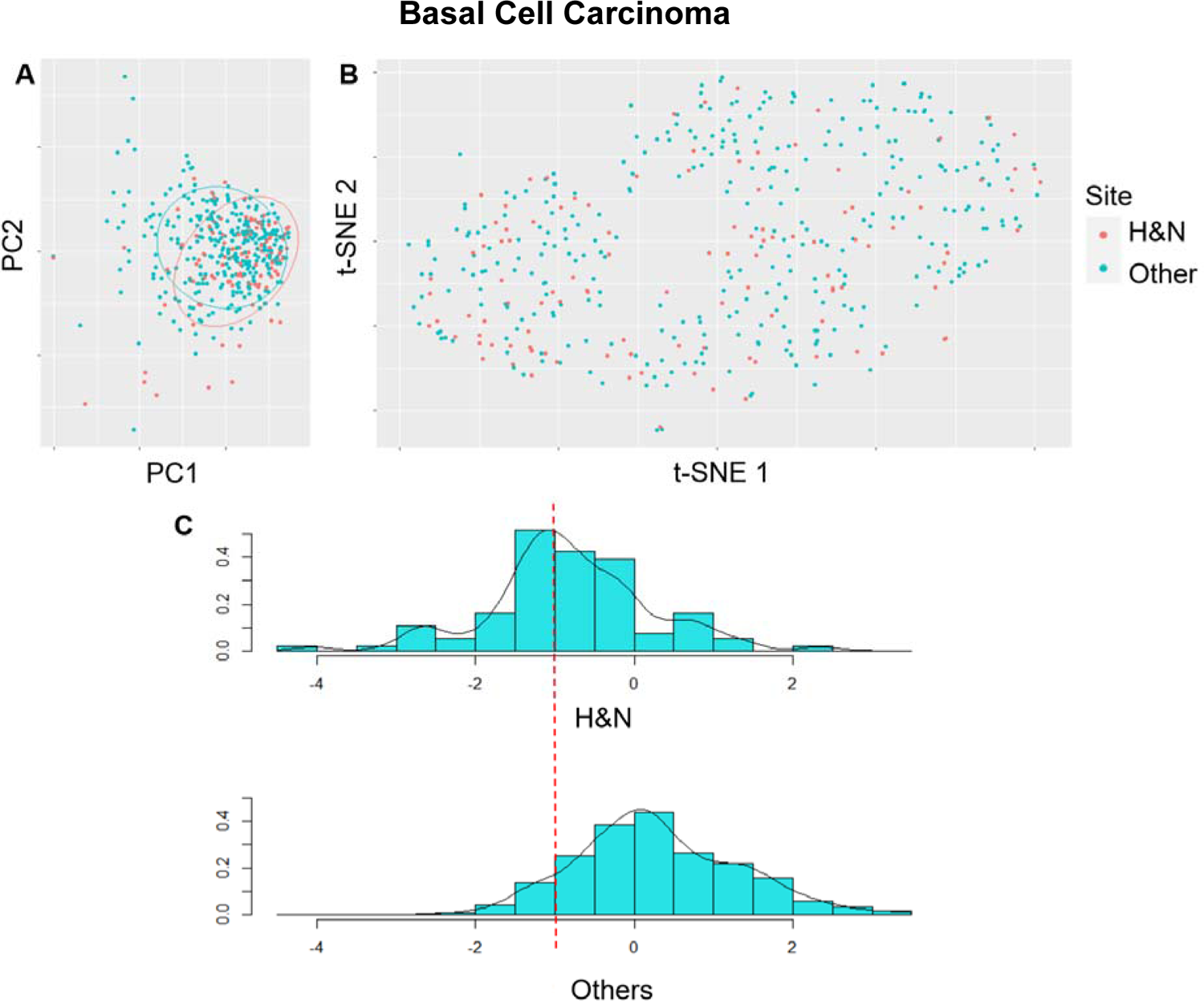
Dimensionality reduction methods employed to further interrogate the BCC multivariate data. (A) Principal component analysis. (B) t-distributed Stochastic Neighbour Embedding plot. (C) Linear discriminant analysis histogram.

The t-SNE plot resulted in clustering of the data into a few groups. While it did not clearly separate H&N and other site BCCs, some clusters appear to have a higher proportion of each anatomic group. For example, a cluster on the left of the t-SNE plot shows a higher proportion of H&N BCCs than the other groups.

### Melanomas of the head & neck are morphologically homogenous

Univariate analysis revealed 13 statistically significant variables (Table 4). H&N melanomas were more common in males than females, *X*^2^(1, n=868) =12.495, *P*<0.001. Eight morphometric features were statistically significantly different on multivariable logistic regression between H&N melanomas and other sites (Table 5). However, the McFadden R^2^ of 0.169 did not show an excellent fit of the model. These results revealed that H&N melanomas had more irregular borders with higher compactness (*P*=0.001) and lower solidity (*P*<0.001) (Figure 3).

**Figure 3.**
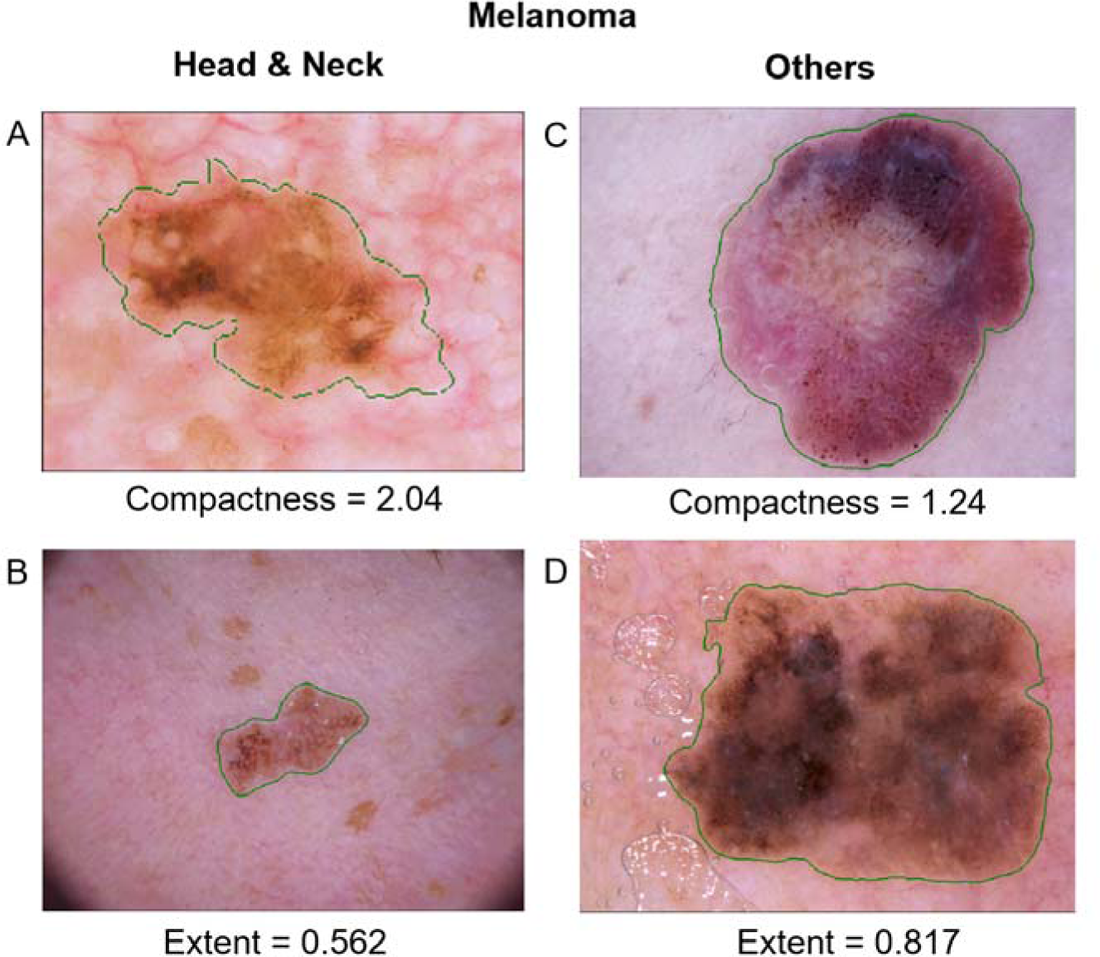
Example melanomas of the H&N exhibiting quantifiable morphological differences to melanomas of other body sites. (A) and (B) showing melanomas of the H&N with higher compactness and lower extent compared to melanomas of other sites, (C) and (D).

**Table 4.** Statistically significant melanoma features on univariate Wilcoxon’s rank sum test. Statistical significance accepted at *P*<0.05.

| | H&N<br>Median | H&N<br>IQR | Others<br>Median | Others<br>IQR | Univariate<br>Wilcoxon's<br>Rank Sum<br>Test $P$ |
| --- | --- | --- | --- | --- | --- |
| Form Factor | 0.717 | 0.113 | 0.751 | 0.119 | 0.001 |
| Compactness | 1.39 | 0.22 | 1.33 | 0.22 | 0.001 |
| Extent | 0.697 | 0.074 | 0.711 | 0.071 | 0.015 |
| Hu Moment 2 | 1.46E-04 | 2.31E-04 | 1.22E-04 | 2.21E-04 | 0.017 |
| Normalized<br>Moment 2,2 | 5.20E-03 | 1.09E-03 | 4.94E-03 | 1.08E-03 | 0.011 |
| Solidity | 0.947 | 0.041 | 0.955 | 0.038 | <0.001 |
| Zernike 3,3 | 1.73E-02 | 1.55E-02 | 1.60E-02 | 1.45E-02 | 0.019 |
| Zernike 6,2 | 1.35E-02 | 9.92E-03 | 1.50E-02 | 9.88E-03 | 0.015 |
| Zernike 7,7 | 6.07E-03 | 5.53E-03 | 5.08E-03 | 4.35E-03 | 0.006 |
| Zernike 8,0 | 7.00E-03 | 6.74E-03 | 5.77E-03 | 6.16E-03 | 0.042 |
| Zernike 8,2 | 6.62E-03 | 5.37E-03 | 7.46E-03 | 5.47E-03 | 0.048 |
| Age | 65.0 | 20.0 | 60.0 | 20.0 | 0.003 |

**Table 5.** Statistically significant melanoma features on multivariable logistic regression. Statistical significance accepted at *P*<0.05.

|  | **H&N**<br>**Median** | **H&N**<br>**IQR** | **Others**<br>**Median** | **Others**<br>**IQR** | **Multivariable**<br>**Logistic**<br>**Regression**<br>***P*** |
| --- | --- | --- | --- | --- | --- |
| Central<br>Moment 0,2 | 3.52E+08 | 7.13E+08 | 4.05E+08 | 8.71E+08 | 0.001 |
| Central<br>Moment 0,3 | -1.61E+07 | 3.88E+09 | 6.97E+06 | 2.66E+09 | 0.016 |
| Inertia Tensor<br>0,0 | 5606 | 5972 | 6133 | 6865 | 0.050 |
| Spatial Moment<br>1,0 | 9.04E+06 | 1.19E+07 | 9.05E+06 | 1.36E+07 | 0.046 |
| Spatial Moment<br>1,1 | 1.40E+09 | 2.66E+09 | 1.49E+09 | 2.98E+09 | 0.029 |
| Spatial Moment<br>2,0 | 1.63E+09 | 2.98E+09 | 1.56E+09 | 3.31E+09 | 0.044 |
| Spatial Moment<br>2,1 | 2.60E+11 | 5.43E+11 | 2.53E+11 | 6.86E+11 | 0.037 |
| Zernike 7,7 | 6.07E-03 | 5.53E-03 | 5.08E-03 | 4.35E-03 | 0.025 |

There was limited separation between the H&N and other site group on the various dimensionality reduction methods (Figure 4). This suggests that while there are some quantifiable morphometric differences between the two anatomical groups, melanomas are overall morphologically homogenous.

**Figure 4.**
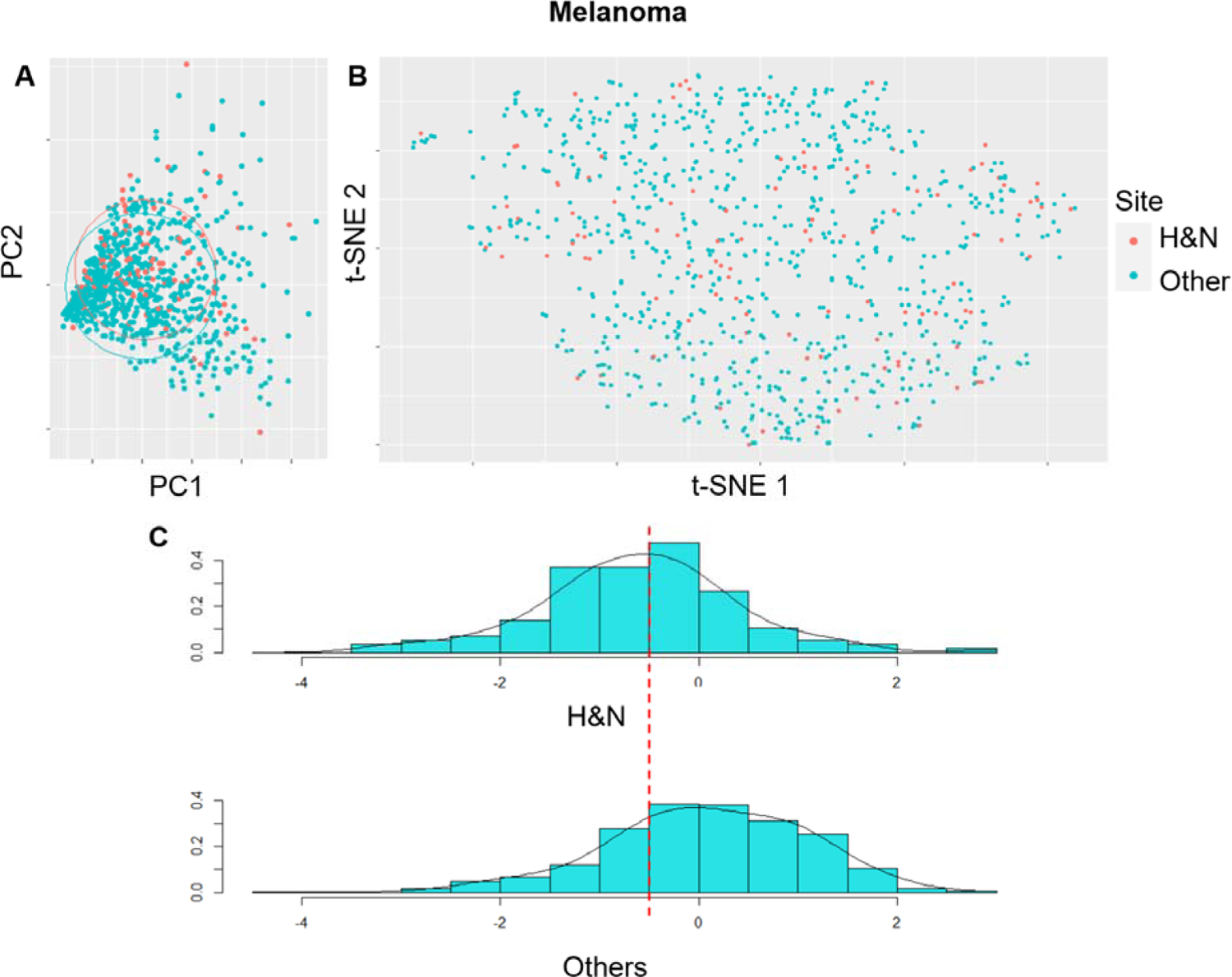
Dimensionality reduction methods employed to further interrogate the melanoma multivariate data. (A) Principal component analysis. (B) t-distributed Stochastic Neighbour Embedding plot. (C) Linear discriminant analysis histogram.

## Discussion

In this study, we have identified several quantifiable morphological differences between BCCs and melanomas of the H&N compared to their respective lesions of other anatomical sites. Unsurprisingly, upon further interrogation of the data, we found substantial overlap between the two sites for both BCCs and melanomas. This is expected as they are pathologically the same lesion and we would expect many of them to look similar dermoscopically. Melanomas exhibited greater homogeneity across their different anatomical locations with little separation on the various data visualisation and dimensionality reduction tools. On the other hand, BCCs showed moderate separation with dimensionality reduction using LDA. This may allude to a clinically significant difference in morphological appearance of H&N BCCs.

Although there was no distinct separation of the BCC data on the t-SNE plot by anatomic site, the data did separate into visible clusters. One cluster on the left of the BCC t-SNE had a greater proportion of H&N BCCs while other clusters at the top showed greater proportions of BCCs from other sites. This may be a result of the data clustering based on BCC subtypes which exhibit different dermoscopic features. Certain BCC subtypes such as the nodular BCC are more common in the H&N region^16,17^ which may explain the observed clustering patterns on the t-SNE plot. However, the HAM10000 dataset does not have information on the BCC subtypes and further studies will be required to investigate this.

While several gene mutations have been identified in the pathogenesis of BCCs, they remain a heterogeneous malignancy with several subtypes that have different clinical behaviours. Furthermore, certain subtypes of BCC have a predilection to specific anatomical sites^16,18^. Anatomic location also influences the likelihood of sun exposure and may contribute to such observed variations. The impact of the underlying anatomy such as vasculature and local lymphatic system has not been thoroughly investigated for their influence on the pathogenesis of BCCs and melanomas. In recent years, there have been advancements in various *in silico, in vitro* and *in vivo* techniques such as vasculature-on-chip devices^19^ and computational models^20,21^ which may enable us to test some of these hypotheses. Further investigations such as computational modelling of skin cancer progression and cancer-on-chip are still required to elucidate the underlying mechanisms to explain the observed heterogeneity of these tumours. However, deriving the mechanism for the observed differences is beyond the scope of this study.

### Implication on AI model diagnoses

In recent years, several AI models for skin lesion classification have been developed and published in the literature. Given the promising results of AI classifiers, we may see their implementation in everyday clinical practice in the near future. In theory, these measurable differences may skew the accuracy of AI classifiers that utilise morphological features or derivatives of such features when generating feature maps for training. The impact of morphological differences will also depend on the training data and its proportion of lesions from various anatomical sites. A model trained on a larger proportion of non-H&N BCCs may result in poorer accuracies when faced with BCCs of the H&N. The influence of these morphometric differences may not be easily identified due to the black box nature of many deep learning models. Therefore, we propose the reporting of site-specific accuracy metrics with complete and accurate metadata in future prospective studies to overcome this issue, especially in the case of BCCs. This will ensure models are generalisable and equally useful to all clinician end-users.

This is particularly important in the H&N region as it represents a unique location that has implications on its management. Speciality clinics such as oral and maxillofacial surgeons, facial plastic surgeons or H&N surgeons may only see skin lesions of the H&N region. Therefore, if AI applications are used in these clinics, the classification accuracy may consistently be skewed and differ from the reported metrics in validation studies. When developing applications and tools, it is imperative to consider the end-users. Therefore, future AI applications for skin cancer may benefit from having dedicated models optimised for use in the H&N to serve this unique anatomical site.

### Implication on AI model training

The findings of this study also has implications on AI model development. For an AI diagnostic model to be robust, it must be trained on data that adequately encompasses the heterogeneity of the condition. Despite the anatomic morphological differences, AI models can still be accurate if they are trained on sufficient H&N dermoscopic images.

Several AI models are trained and validated on images from public datasets of dermoscopic images. There are several dermoscopic datasets that are freely available such as the HAM10000, used in this study, and the PH2 dataset. In these two examples, the images were acquired from a dermatology clinic which may not be representative of the full anatomic range of skin cancers. Some BCCs and melanomas of the H&N may have presented to other speciality departments such as oral and maxillofacial surgery and plastic surgery. The under-representation of these skin cancers from the H&N sites in these datasets would in turn influence AI models trained on these datasets, potentially making these models less accurate in the H&N sites.

When comparing the proportion of H&N BCCs and melanomas in the HAM10000 dataset to other published studies, we can see there is indeed a smaller proportion of BCCs and melanomas in the HAM10000 dataset. In this study, 26.5% of BCCs and 13.0% of melanomas came from the H&N region. However, the H&N is a common location for BCCs^22^ and melanomas^23^. Other studies with much larger cohorts from state or nation-wide cancer registries report up to 40% of BCCs^22^ and approximately 20-30% of melanomas arising in the H&N^23,24^.

This highlights the significant under-representation of the H&N skin cancers from public datasets collected from dermatology departments. Furthermore, with the morphological differences between H&N and other body sites, these factors may result in consistently poorer accuracies if AI models were employed by the H&N specialities managing skin cancers. These reasons support the need for more thorough validation of AI models or even dedicated AI models for use on H&N skin cancers.

For clinicians and the potential end-users of AI models for diagnosing skin lesions, we must also scrutinise the methods employed to develop the AI models we are using to ensure their training data is representative and can be generalised for use in our own clinics and patient populations.

### Limitations

It should be noted that some of the morphological measurements such as spatial and central moments make use of the objects’ size alongside their shape and orientation. However, there is no information on whether the dermoscopic images from the HAM10000 dataset have been calibrated for size. As they are likely not calibrated, the distance at which the lesion was imaged and the optical magnification will influence the size of the imaged lesion. The authors of the original paper reported no post-processing apart from centering and cropping edges of the dermoscopic images.

All the HAM10000 dermoscopic images were acquired via contact dermoscopy which ensures the distance from dermoscope to lesion is relatively constant. The issue of pixel spacing is more of a problem with non-contact dermoscopy where the distance from dermoscope to lesion can be varied. Furthermore, even without calibration, majority of the HAM10000 images were acquired with contact dermoscopes of 10x optical magnification with the exception of those acquired with the Molemax HD system (optical magnification up to 100x) and the Heine Delta 20 (optical magnification from 10-16x). Several spatial and central moments were found to be significantly different between the two anatomical groups. We opted to leave these features in our analysis as they also encode other useful morphological information and the majority of the images would be comparable in terms of pixel spacing even without calibration. The intention of this study is to reveal potential morphological differences between anatomical sites to caution specialist clinicians operating only in specific anatomical areas on the potential influence this may have on AI diagnostic models. Ideally, these results should be confirmed with appropriately calibrated dermoscopic images with the same pixel spacing.

In addition, Hu moments and normalized moment features are not influenced by the objects’ size or location and several of these features were also found to be statistically significant.

## Conclusion

In conclusion, we have identified several morphological differences between BCCs and melanomas of the H&N region when compared to other body sites. Overall, melanomas are more homogenous across different anatomical sites but BCCs may exhibit greater morphometric variation. These findings support the need for site-specific validation of image-based diagnostic AI models for skin cancers. Furthermore, when considering the clinician end-users and these morphologic differences, there may be utility in developing dedicated tools for H&N skin cancer diagnostics.

## Data Availability

All data produced in the present study are available upon reasonable request to the authors.

## Acknowledgements

This research did not receive any specific grant from funding agencies in the public, commercial, or not-for-profit sectors.

